# Accelerometry-derived activity fragmentation as a predictor of brain atrophy and disability progression in multiple sclerosis

**DOI:** 10.1101/2025.09.02.25334901

**Authors:** Angeliki G Filippatou, Muraleetharan Sanjayan, Blake Dewey, Evan Johnson, Pratim Guha Niyogi, Yasser Fadlallah, Safiya Duncan, Chelsea Wyche, Ela Moni, Vadim Zipunnikov, Ellen M Mowry, Kathryn C Fitzgerald

## Abstract

**Background:** Accelerometry-derived activity fragmentation reflects how frequently active periods are broken by sedentary periods. It has been associated with frailty, mortality, and reduced gait speed in the general population; however, its prognostic significance in multiple sclerosis (MS) remains unknown.

**Methods:** People with MS (PwMS) wore wrist tri-axial accelerometers at three-month intervals, underwent disability assessments biannually and MRI brain approximately annually. We evaluated whether active-to-sedentary transition probability (ASTP) and sedentary-to-active transition probability (SATP) during the 10 most active hours of each 24-hour period (M10) were associated with confirmed disability progression and brain substructure atrophy, by modeling between- and within-person effects.

**Results:** Among 239 PwMS, 120 had confirmed Expanded Disability Status Scale(EDSS)-plus progression over a mean of 2.9±1.1 years. Within-person increase in ASTP slope during M10 (indicating increasing activity fragmentation throughout M10) was associated with higher risk of subsequent EDSS-plus progression (per 1 SD increase: HR 1.22 [95%CI 1.02,1.46],p=0.03) and lower deep gray matter and thalamic volumes over time (per 1 SD increase: -0.35 [95%CI - 0.58,-0.13],p=0.002, -0.42 [95%CI:-0.68,-0.15],p=0.002 respectively). There were no similar associations for SATP.

**Conclusions:** Within-person worsening in activity fragmentation was associated with higher risk of neurologic decline. Activity fragmentation may represent an early compensatory strategy for reduced physiologic reserve and serve as an early indicator of MS progression.

## INTRODUCTION

Multiple sclerosis (MS) is a chronic neuroinflammatory and neurodegenerative disorder of the central nervous system.^1^ Throughout their disease course, people with MS (PwMS) may experience relapses that are caused by immune-mediated demyelination, and progression that is characterized by gradual deterioration of function and is caused by neurodegeneration.^2^ Therapeutic development for progressive stages of MS has been hampered by the lack of sensitive measures of disease progression, particularly over short timeframes.^3-5^ Wearable biosensors, such as accelerometers, have emerged as promising tools for more precise assessment of functional status in PwMS.^6-9^

Accelerometers provide objective, real-world data across multiple dimensions of movement by capturing general patterns of physical activity and sleep as well as more subtle behavioral patterns like activity fragmentation. Activity fragmentation refers to the degree to which activity is interrupted by transitions between active and sedentary states (**Figure 1**). Higher active state fragmentation (more fragmented periods of activity) has been linked to functional limitations, frailty and higher mortality risk in the general population.^10-17^ Active state fragmentation has also been associated with fatigability and slower gait speed in older adults; it is hypothesized to represent an early compensatory strategy in the context of diminished stamina and physiologic decline.^14,18,19^ Conversely, higher sedentary state fragmentation (more fragmented sedentary time) is hypothesized to reflect better cardiometabolic health due to promotion of increased energy expenditure.^15^ Measures of activity fragmentation may be relevant in MS, since PwMS are particularly susceptible to fatigability and diminished endurance, which may lead to progressive shortening of activity bouts that can be tolerated without resting as the day progresses. A prior cross-sectional study in PwMS found differences in measures of activity fragmentation compared to healthy controls, indicating that PwMS exhibit more fragmented active behavior.^20^ Moreover, in a recent paper reporting the baseline data from our longitudinal observational study Home-based Evaluation of Actigraphy to predict Longitudinal Function in MS (HEAL-MS), we found that individuals with progressive MS (PMS) had higher activity fragmentation compared to those with relapsing-remitting MS (RRMS).^21^ However, longitudinal studies assessing how changes in activity fragmentation over the course of a given day and over time relate to subsequent disability progression are lacking.

**Figure 1.**
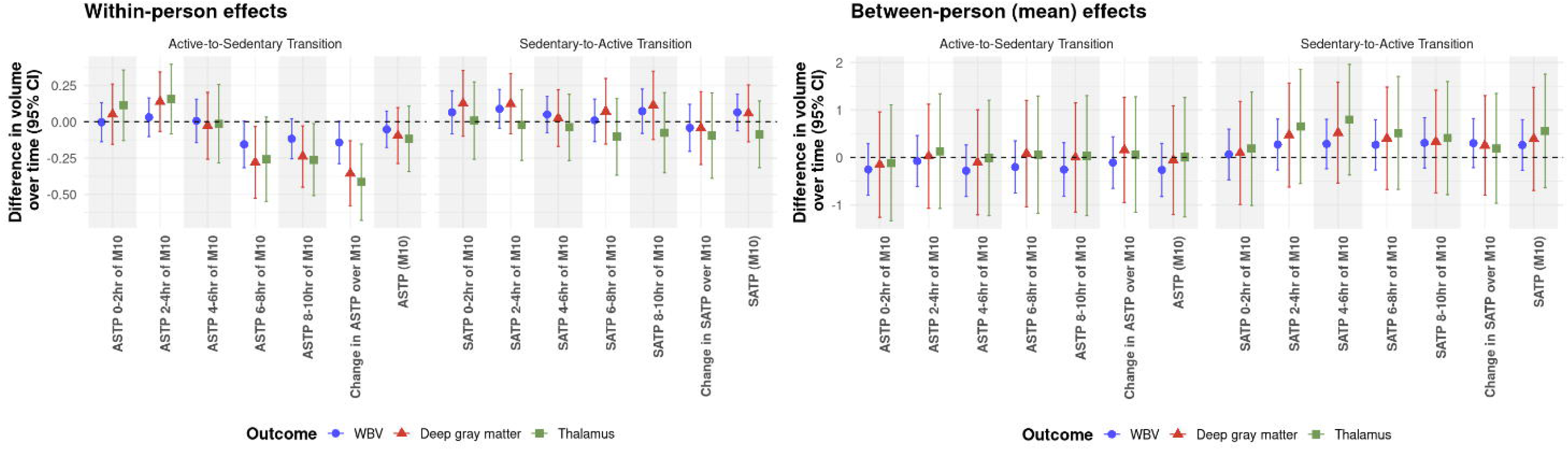
Activity fragmentation conceptualization. The top row shows an individual performing continuous activity followed by sedentary time. The bottom row shows an individual with alternating active and sedentary intervals, breaking up their activity into shorter, more fragmented bouts. *Figure created in BioRender.* https://BioRender.com/vqnld5o

In this longitudinal study of the HEAL-MS cohort, we sought to evaluate how accelerometry-derived fragmentation metrics change over time and, if within-person changes in activity fragmentation during follow-up are associated with brain compartment atrophy rates and disability progression. We hypothesized that within-person worsening in activity fragmentation throughout the course of the day is a proxy of fatigability and reduced physiologic reserve, and that it would be associated with downstream accelerated brain atrophy and disability progression during follow-up. With regards to brain atrophy, we focused on whole brain as well as deep gray matter (GM), since deep GM atrophy is relevant in MS; it occurs early in the disease course, often precedes measurable whole brain volume loss, and exhibits strong longitudinal associations with disability and cognition.^22^

## METHODS

### Study population

PwMS were recruited from the Johns Hopkins MS Center into the HEAL-MS study as previously described.^21^ Briefly, HEAL-MS participants were recruited between January 2021 and March 2023. Eligibility criteria included MS diagnosis based on 2017 McDonald diagnostic criteria^23^, age ≥ 40 years, EDSS ≤ 6.5, no relapse in the six months prior to enrollment, and absence of identified comorbidities that may limit physical activity. The inclusion criteria were selected to ensure the cohort included PwMS who already had, or were at risk (based on age) of transitioning to, PMS, to avoid confounding of baseline scores by incomplete recovery from recent relapse, and to ensure that changes in accelerometry were likely to be related to MS rather than comorbid conditions. By design, one-third of the cohort had PMS at baseline, while the remainder were classified as RRMS by an MS-trained neurologist. Participants with RRMS were further classified into stable RRMS (with no suspected or confirmed disability progression) and RRMS with suspected clinical progression but no sustained disability worsening at study entry, per the clinical judgment of a neurologist on the study team.

### Accelerometry measures

HEAL-MS participants wore GT9X Actigraph wrist tri-axial accelerometers continuously over a two-week period at pre-specified, three-month intervals. Participants were instructed to wear the device on the wrist of their non-dominant hand continuously for 24 hours a day over the duration of two weeks for each wear period. Accelerometers were set to capture three-dimensional acceleration at 30 Hz with a range of ± 8 G. Accelerometry data were processed as previously described.^21^ Briefly, raw acceleration data were converted into 60-second epoch vector magnitude-based activity count data. Non-wear intervals were defined as intervals of 90 minutes or longer in which all minute-level activity counts were zero.^21^ Valid days during each wear period were defined as those in which the total wear time was at least 90% of the day. We excluded participants with fewer than three valid wear days per wear period.

Accelerometry measures of activity fragmentation included the active to sedentary transition probability (ASTP) and sedentary to active transition probability (SATP), which were calculated as previously described.^21,24^ An active minute was defined as a minute with activity counts > 2000 and a sedentary minute was defined as a minute with activity counts ≤2000. We subsequently calculated the average length of active and sedentary bouts. ASPT represents the inverse of the average length of active bouts, and SAPT represents the inverse of the average length of sedentary bouts. Higher ASTP values imply higher active state fragmentation, i.e. more fragmented or less sustained periods of activity, while higher SATP values imply higher sedentary state fragmentation, i.e. more fragmented or less sustained periods of sedentary time.

We calculated the following formulations of ASTP and SATP reflecting overall fragmentation, as well as how fragmentation changes over the course of a day:

1. Mean ASTP and SATP during a person’s most active 10 hours of a 24-hour period (M10)
2. Mean ASTP and SATP during consecutive two-hour intervals across the M10 period (i.e. mean ASTP or SATP during 0-2h, 2-4h, 4-6h, 6-8h, 8-10h of M10). Consistent with prior work ^8,21^, two-hour intervals offer greater resolution in characterizing activity patterns during M10.
3. Intra-day change in ASPT and SAPT throughout M10; quantified as the linear slope of each metric over M10 (i.e., a regression line considering how ASTP changes across 0-2h, 2-4h, 4-6h, 6-8h, 8-10h during M10)

### Clinical measures

HEAL-MS participants underwent study evaluations every six months through scheduled visits. At each visit, an EDSS was conducted by a masked EDSS-trained physician, and a modified MS Functional Composite (MSFC) was conducted by a masked, trained study team member. The modified MSFC includes a 9-hole peg test (9HPT), timed 25-foot walk test (T25FW), binocular 2.5% contrast visual acuity test, and the Symbol Digit Modalities Test (SDMT) cognitive assessment. Whenever possible, to minimize variance introduced by transient physical fatigue (e.g. due to walking into clinic), disability assessments were conducted at least 15 minutes after the patient arrived and was seated.

The primary clinical outcome for disability for our study was the EDSS-plus, a composite that includes confirmed EDSS progression (defined as change by >1.0 point if baseline EDSS < 5.5 or > 0.5 if baseline EDSS ≥ 6.0) OR 20% worsening on either of two specific components of the MSFC (T25FW or 9HPT), that is sustained ≥ 24 weeks later. We considered a roving baseline, which has been reported to increase sensitivity in detecting disability progression.^25^

### Imaging measures

Brain MRI scans were acquired on a 3T Siemens scanner at baseline and end of year two of follow-up as part of the HEAL-MS protocol. Additional MRIs acquired under an identical protocol as part of clinical care were also included when available. As part of the MRI processing pipeline, multi-slice image volumes undergo deep learning-based super-resolution with Synthetic Multi-Orientation Resolution Enhancement (SMORE), followed by alignement of the super-resolved and 3D acquired images using a longitudinal registration system.^26-28^ As part of the alignement process, each subject is aligned to a reference atlas, images are adjusted across different time points and within each time point. Rigid transformations are employed to maintain anatomical integrity. After Harmonization with Attention-based Contrast, Anatomy and Artifact Awareness (HACA3), three contrast targets are generated at each time point: 3D magnetization-prepared rapid gradient-echo (MPRAGE), 3D fluid-attenuated inversion recovery (FLAIR;), multi-slice T2-weighted.^29^ Lesion segmentation was performed using an in-house deep-learning model based on Self-Ensembled Lesion Fusion.^30^ Brain substructure volumes were generated after lesion filling by utilizing Spatially Localized Atlas Network Tiles (SLANT).^31,32^ After segmentation, the surface is reconstructed using Cortical Reconstruction Using Implicit Surface Evolution (CRUISE) to refine SLANT segmentation boundaries and measure cortical thickness.^33,34^ A deep neural network was used to calculate the total intracranial volume (ICV); all MRI volumes were normalized for ICV to obtain volume fractions, thus accounting for variation in head size.^35^

### Statistical methods

Statistical analyses were conducted using R. Baseline characteristics of the cohort are summarized using descriptive statistics (mean ± standard deviation [SD] or median with interquartile range [IQR]) where applicable. Firstly, we estimated changes in activity fragmentation metrics during follow-up using linear mixed effects models adjusted for age, sex, race and body mass index (BMI), to assess how activity fragmentation evolves in PwMS evolves. Subsequently, to understand how these changes translate to outcomes, we modeled within-person effects to evaluate whether changes in fragmentation metrics were associated with brain compartment atrophy rates and time to confirmed disability progression at the individual level.^36,37^ This analytic structure allows us to evaluate both between-person (average or group-level) effects and within-person (individual) effects. For this analysis, we calculated an overall person-specific mean of each fragmentation metric across all visits, and a within-person measure, calculated as the difference between the overall person-specific mean of the fragmentation metric and the fragmentation metric at each wear period. For each outcome, we fit a regression model that included both the overall mean of a given metric (between-person effect) and the within-person measure, with both terms standardized to reflect a 1 SD unit difference. For disability outcomes, Cox regression considering time-varying fragmentation measures was used to analyze time to confirmed disability progression, with models additionally adjusted for age, sex, race and BMI. Only accelerometry measures collected prior to the event of confirmed disability progression were included for quantifying person-specific means. For MRI outcomes, linear regression was used, and models were adjusted for a similar set of covariates with the addition of EDSS. Sensitivity analyses were additionally adjusted for accelerometry-derived total activity count (TAC) to ensure that any observed associations were not reflective of differences in overall activity levels.

### Ethical Considerations

The study was approved by the Institutional Review Board of the Johns Hopkins University. All participants provided written informed consent prior to study enrolment.

## RESULTS

### Cohort description

A total of 239 PwMS were included in this study (mean age 54.7±8.6, 71.1% female, 23% non-white). At baseline, 81 participants were classified as PMS, 79 as stable RRMS, and 79 as RRMS with suspected progression. Median baseline EDSS score was 3. Baseline characteristics for the overall cohort and by HEAL-MS subgroup are presented in **Table 1**. Participants were followed for an average of 2.9 years for the disability outcomes and 1.2 years for the MRI outcomes.

**Table 1.** Demographic characteristics of HEAL-MS participants.

|  | Overall | Relapsing MS (stable) | Relapsing MS (suspected progression) | Progressive MS |
| --- | --- | --- | --- | --- |
| N | 239 | 79 | 79 | 81 |
| Age, mean (SD), years | 54.7 (8.6) | 52.8 (7.5) | 53.8 (8.4) | 57.5 (9.3) |
| Male sex, n (%) | 69 (28.9) | 24 (30.4) | 19 (24.1) | 26 (32.1) |
| Race, n (%) |  |  |  |  |
| White | 184 (77.0) | 60 (75.9) | 60 (75.9) | 64 (79.0) |
| Black | 42 (17.6) | 17 (21.5) | 13 (16.5) | 12 (14.8) |
| Other | 7 (2.9) | 0 (0.0) | 2 (2.5) | 5 (6.2) |
| Hispanic or Latinx ethnicity, n (%) | 9 (4.0) | 3 (3.9) | 3 (4.2) | 3 (3.8) |
| Number of accelerometry wears, mean (SD) | 9.61 (3.97) | 9.59 (3.81) | 9.42 (4.27) | 9.80 (3.85) |
| Total number of days of accelerometry wear, mean (SD) | 86.07 (38.44) | 86.92 (37.20) | 84.20 (41.25) | 87.05 (37.18) |
| Number of days per accelerometry wear, mean (SD) | 8.84 (1.07) | 8.91 (1.18) | 8.81 (1.09) | 8.80 (0.93) |
| N EDSS/MSFC assessments, mean (SD) | 6.03 (2.04) | 6.30 (1.69) | 5.92 (2.18) | 5.85 (2.21) |
| Disability outcome follow-up time, mean (SD), years | 2.89 (1.09) | 2.97 (0.74) | 2.94 (1.36) | 2.75 (1.07) |
| N MRIs, mean (SD) | 2.22 (1.02) | 2.20 (0.94) | 2.33 (1.09) | 2.14 (1.02) |
| MRI Follow-up time, mean (SD), years | 1.17 (0.93) | 1.13 (0.91) | 1.21 (0.96) | 1.16 (0.94) |
| Body mass index, mean (SD), kg/m<sup>2</sup> | 28.60 (6.10) | 29.19 (6.11) | 28.40 (5.90) | 28.20 (6.30) |
| MS symptom duration, mean (SD), years | 13.24 (8.21) | 11.75 (7.22) | 13.79 (8.03) | 14.11 (9.12) |
| MS DMT, n (%) |  |  |  |  |
| Injectable interferon or glatiramer | 41 (17.2) | 12 (15.2) | 15 (19.0) | 14 (17.3) |
| Oral | 39 (16.3) | 19 (24.1) | 15 (19.0) | 5 (6.2) |
| Infusion or immune reconstitution | 103 (43.1) | 32 (40.5) | 39 (49.4) | 32 (39.5) |
| None | 56 (23.4) | 16 (20.3) | 10 (12.7) | 30 (37.0) |
| Baseline EDSS, median (IQR) | 3 (2.0, 4.5) | 2 (1.75-3.0) | 3 (2.0-3.5) | 5 (3.5-6.5) |

MS: multiple sclerosis; SD: standard deviation; IQR: interquartile range, EDSS: expanded disability status scale; MSFC: MS functional composite; DMT: disease-modifying treatment

### Changes in activity fragmentation

Participants wore accelerometers an average of 9.6 times, reflecting a total of 86 days of wear during a mean follow-up period of 2.9 years. ASTP largely increased over time, indicating increasing active state fragmentation during follow-up (more fragmented or less sustained periods of activity), while SATP largely decreased over time, indicating lower sedentary state fragmentation (less fragmented or more sustained periods of sedentary time) **(Figure 2)**. Analyses were consistent when adjusting for season **(Supplementary Figure 1)**. When examining how fragmentation indices changed by MS subtype, these findings were mainly driven by the PMS group **(Supplementary Figure 2)**.

**Figure 2.**
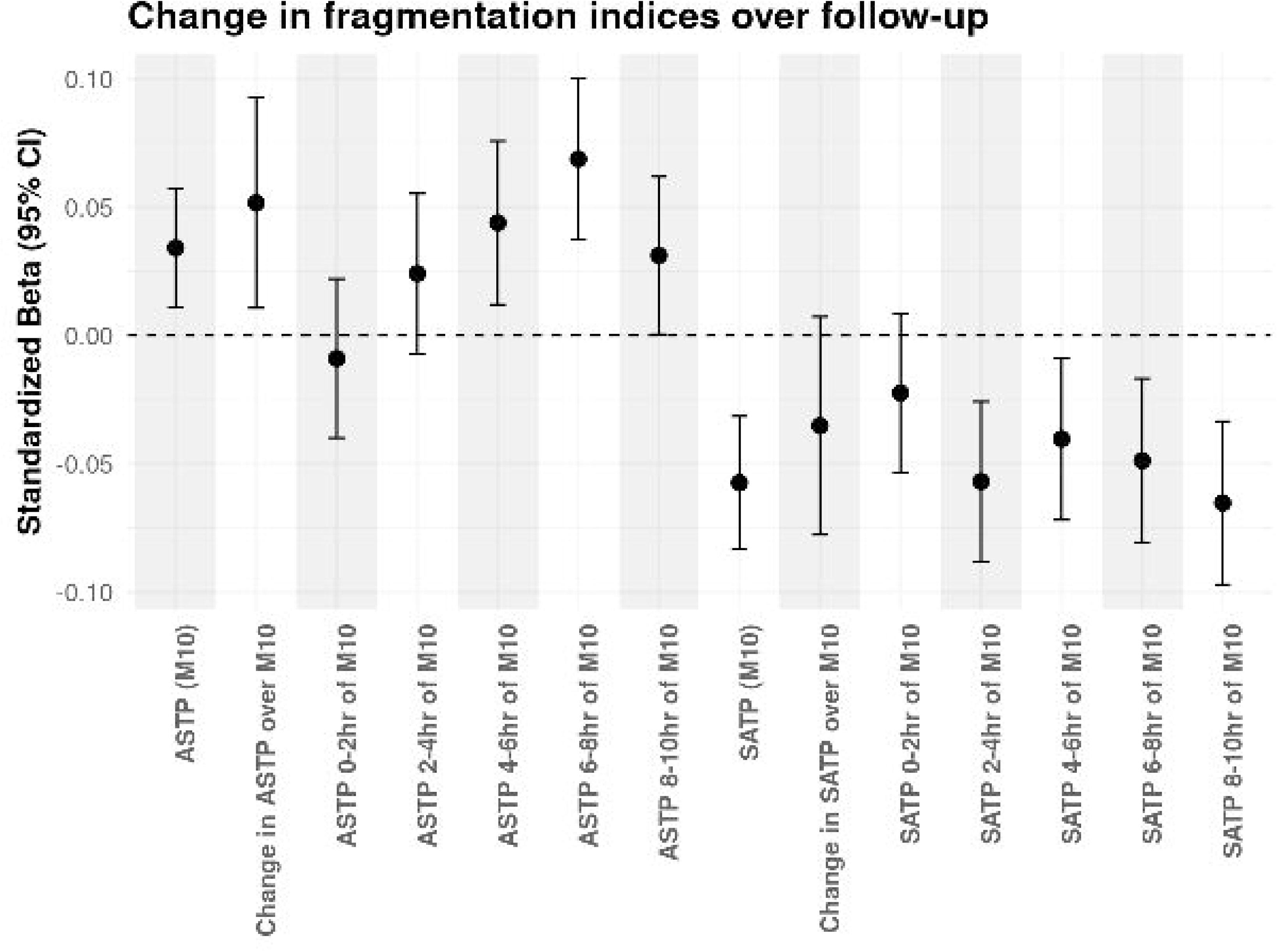
Change in metrics of activity fragmentation during follow-up. *Estimates are calculated from linear mixed effects models adjusted for age, sex, race and body mass index (BMI). Abbreviations: RRMS: relapsing-remitting MS, PMS: progressive MS, ASTP: active-to-sedentary transition probability, SATP: sedentary-to-active transition probability; M10: most active 10 hours of the day*

### Association of activity fragmentation with disability progression

A total of 120 PwMS experienced EDSS-plus progression, sustained 24 weeks later, during follow-up. Within-person increase in ASTP slope during M10 (indicating increasing activity fragmentation throughout the course of M10) was associated with higher risk of confirmed EDSS-plus progression (per 1 SD increase: HR 1.22 [95%CI: 1.02 to 1.46], p=0.03). Within-person increase in ASTP during the final 2 hours of M10 was associated with higher risk of confirmed EDSS-plus progression (per 1 SD increase: HR 1.22 [95%CI: 1.05 to 1.42], p=0.008).

Findings were consistent in analyses for EDSS progression, except within-person increase in average ASTP during M10 was also associated with higher risk of EDSS progression. There were no associations between ASTP and disability progression in mean/between-person models. There were no associations between SATP and disability progression in within-person or between-person models. Findings are summarized in **Table 2** and **Figure 3**. Findings for the primary outcome (EDSS-plus) were similar in sensitivity analyses adjusted for TAC **(Supplementary Table 1)**.

**Figure 3.**
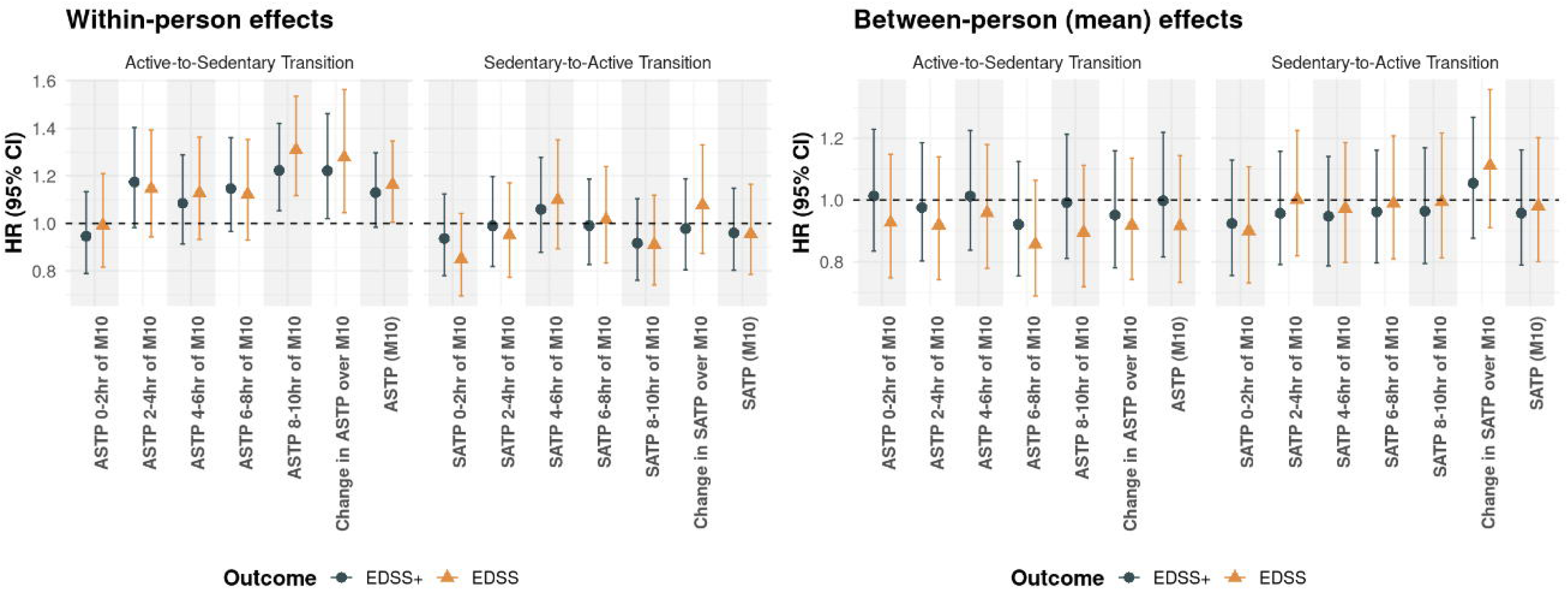
Association of metrics of activity fragmentation with risk of confirmed disability progression during follow-up. *Hazard ratios (HR) are calculated from Cox regression modes adjusted for adjusted for age, sex, race and body mass index (BMI). Each HR reflects the association between a 1 standard deviation (SD) change in activity fragmentation and risk of subsequent disability progression. The reference line at HR=1 indicates no change in risk. The left panel represents within-person effects, and the right panel represents between-person/ mean effects*. *Abbreviations: ASTP: active-to-sedentary transition probability, SATP: sedentary-to-active transition probability; M10: most active 10 hours of the day*

**Table 2.** Association of metrics of activity fragmentation with risk of confirmed disability worsening during follow-up.

| Variable | Confirmed EDSS-plus progression |  |  |  | Confirmed EDSS progression |  |  |  |
| --- | --- | --- | --- | --- | --- | --- | --- | --- |
|  | Mean/between-person model |  | Within-person model |  | Mean/between-person model |  | Within-person model |  |
|  | HR (95 % CI) | P-value | HR (95 % CI) | P-value | HR (95% CI) | P-value | HR (95% CI) | P-value |
| ASTP (M10) | 1.00 (0.82, 1.22) | 0.98 | 1.13 (0.98, 1.30) | 0.08 | 0.92 (0.73, 1.14) | 0.44 | **1.16 (1.00, 1.35)** | **0.04** |
| Change in ASTP over M10 | 0.95 (0.78, 1.16) | 0.62 | **1.22 (1.02, 1.46)** | **0.03** | 0.92 (0.74, 1.14) | 0.43 | **1.28 (1.04, 1.56)** | **0.02** |
| ASTP 0-2hr of M10 | 1.01 (0.83, 1.23) | 0.9 | 0.95 (0.79, 1.13) | 0.55 | 0.93 (0.75, 1.15) | 0.49 | 0.99 (0.82, 1.21) | 0.95 |
| ASTP 2-4hr of M10 | 0.98 (0.80, 1.19) | 0.8 | 1.17 (0.98, 1.40) | 0.08 | 0.92 (0.74, 1.14) | 0.44 | 1.15 (0.94, 1.39) | 0.17 |
| ASTP 4-6hr of M10 | 1.01 (0.84, 1.23) | 0.9 | 1.08 (0.91, 1.29) | 0.35 | 0.96 (0.78, 1.18) | 0.69 | 1.13 (0.93, 1.36) | 0.21 |
| ASTP 6-8hr of M10 | 0.92 (0.75, 1.12) | 0.42 | 1.15 (0.97, 1.36) | 0.12 | 0.86 (0.69, 1.06) | 0.16 | 1.12 (0.93, 1.35) | 0.23 |
| ASTP 8-10hr of M10 | 0.99 (0.81, 1.21) | 0.94 | **1.22 (1.05, 1.42)** | **0.008** | 0.89 (0.72, 1.11) | 0.32 | **1.31 (1.12, 1.54)** | **0.001** |
| SATP (M10) | 0.96 (0.79, 1.16) | 0.66 | 0.96 (0.80, 1.15) | 0.66 | 0.98 (0.80, 1.20) | 0.85 | 0.96 (0.79, 1.16) | 0.66 |
| Change in SATP over M10 | 1.05 (0.88, 1.27) | 0.57 | 0.98 (0.81, 1.19) | 0.82 | 1.11 (0.91, 1.36) | 0.3 | 1.08 (0.87, 1.33) | 0.48 |
| SATP 0-2hr of M10 | 0.92 (0.76, 1.13) | 0.44 | 0.94 (0.78, 1.12) | 0.48 | 0.90 (0.73, 1.11) | 0.32 | 0.85 (0.70, 1.04) | 0.12 |
| SATP 2-4hr of M10 | 0.96 (0.79, 1.16) | 0.65 | 0.99 (0.82, 1.20) | 0.92 | 1.00 (0.82, 1.23) | 0.99 | 0.95 (0.77, 1.17) | 0.64 |
| SATP 4-6hr of M10 | 0.95 (0.79, 1.14) | 0.57 | 1.06 (0.88, 1.28) | 0.55 | 0.97 (0.80, 1.19) | 0.79 | 1.10 (0.89, 1.35) | 0.37 |
| SATP 6-8hr of M10 | 0.96 (0.80, 1.16) | 0.68 | 0.99 (0.83, 1.19) | 0.92 | 0.99 (0.81, 1.21) | 0.91 | 1.02 (0.83, 1.24) | 0.88 |
| SATP 8-10hr of M10 | 0.96 (0.79, 1.17) | 0.71 | 0.92 (0.76, 1.10) | 0.36 | 0.99 (0.81, 1.22) | 0.96 | 0.91 (0.74, 1.12) | 0.37 |

EDSS: expanded disability status scale; HR: hazard ratio; ASTP: active-to-sedentary transition probability, SATP: sedentary-to-active transition probability; M10: most active 10 hours of the day

### Association of activity fragmentation with brain atrophy

Within-person increase in ASTP slope during M10 was associated with lower thalamic volumes over time (per 1 SD increase: -0.42% [95%CI: -0.68 to -0.15], p=0.002) and deep GM volumes over time (per 1 SD increase: -0.35% [95% CI: -0.58 to -0.13], p=0.002). Similarly, within-person increase in ASTP during the final 2 hours of M10 was associated with lower thalamic and deep GM volumes over time (for thalamus: per 1 SD increase: -0.26% [95%CI: -0.51 to -0.01], p=0.04; for deep GM: per 1 SD increase: -0.24% [95% CI: -0.45 to -0.03], p=0.03). Within-person increase in ASTP between the sixth and eighth hours of M10 was also associated with accelerated deep GM atrophy (per 1 SD increase: -0.28% [95% CI: -0.53 to -0.03], p=0.03). Findings for WBV over time showed the same directional trend, although they did not reach statistical significance. There were no associations between ASTP and longitudinal thalamic, deep GM or WBV volumes in mean/between-person models. Similar to disability outcomes, there were no associations between SATP and brain compartment atrophy rates in within-person or between-person models. Findings are summarized in **Table 3** and **Figure 4**. Findings were largely similar in sensitivity analyses adjusted for TAC **(Supplementary Table 2)**.

**Figure 4.**
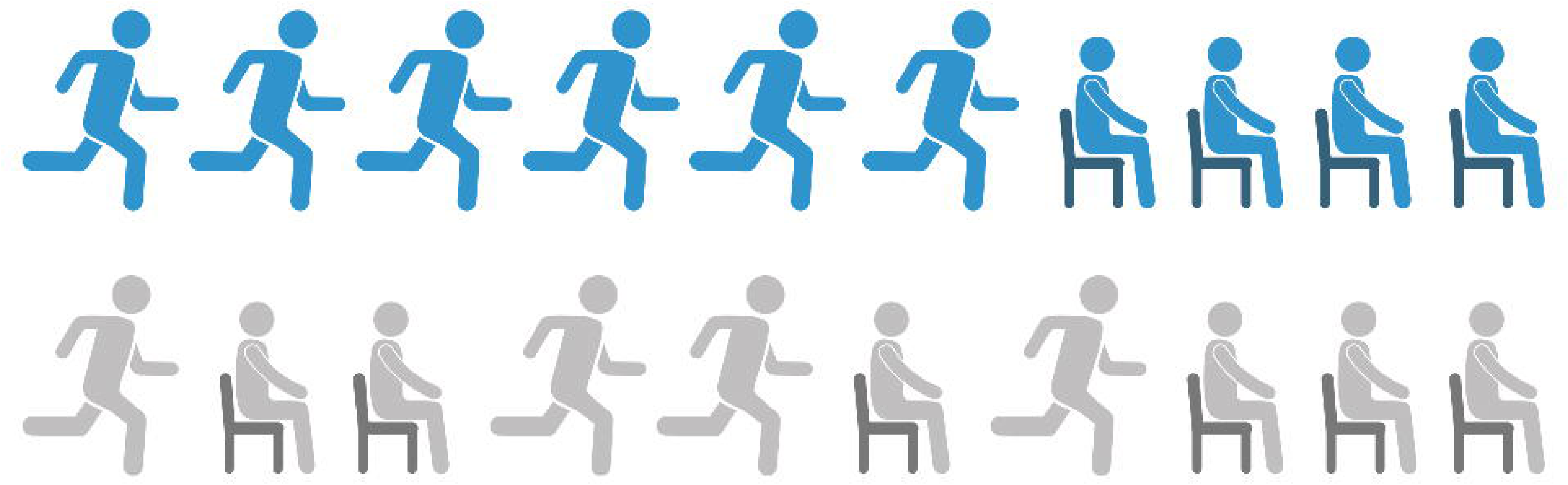
Association of metrics of activity fragmentation with rates of brain compartment atrophy. *Estimates are calculated from linear mixed effects models adjusted for age, sex, race and body mass index (BMI). Each estimate reflects the association between a 1 standard deviation (SD) change in activity fragmentation and difference in brain volume over time. The reference line at 0 indicates no difference in brain volume over time. The left panel represents within-person effects, and the right panel represents between-person/ mean effects*. *Abbreviations: ASTP: active-to-sedentary transition probability, SATP: sedentary-to-active transition probability; M10: most active 10 hours of the day; WBV: whole brain volume*

**Table 3.**
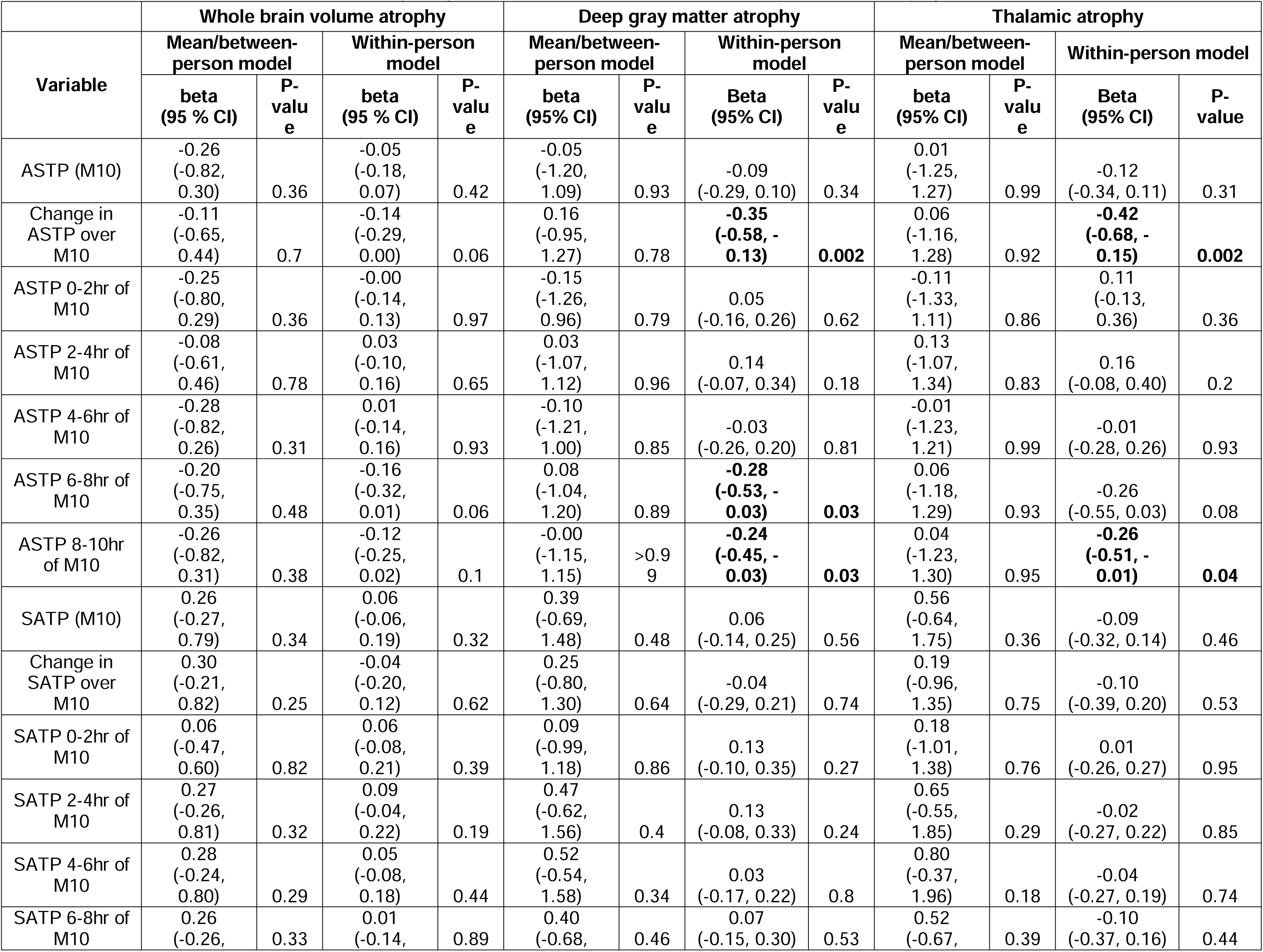
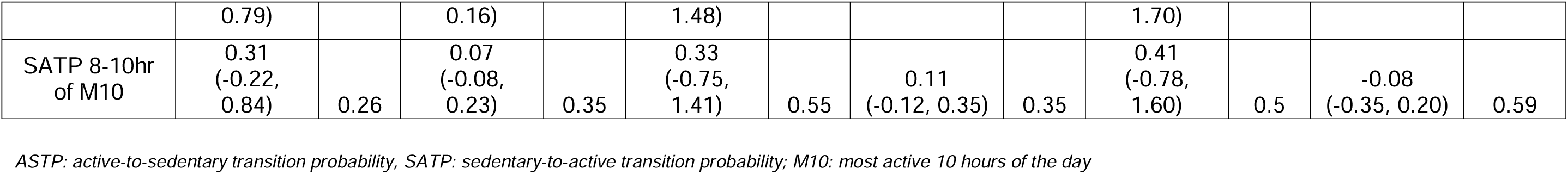
Association of metrics of activity fragmentation with rates of brain compartment atrophy.

## DISCUSSION

In this well-characterized cohort of PwMS that included longitudinal assessments of accelerometry, disability status and brain imaging, an increase in activity fragmentation at the individual level was predictive of disability progression and accelerated brain atrophy. Specifically, we found that ASTP increased over time, and that within-person increase in ASTP during the final hours of M10 and a steeper ASTP slope throughout M10 were associated with higher risk of confirmed disability progression and accelerated thalamic and deep GM atrophy. These longitudinal findings nicely complement our prior cross-sectional results from the same cohort, that demonstrated higher activity fragmentation in individuals with PMS compared to RRMS.^21^ Our findings suggest that PwMS who exhibit worsening activity fragmentation throughout the course of their active period are at a higher risk of subsequent neurological decline. We hypothesize that higher activity fragmentation serves as an early indicator of diminished stamina and reduced physiologic reserve that later manifests as brain atrophy and disability progression.

One noteworthy aspect of our findings is that the observed associations between disability and imaging outcomes and ASTP were particularly driven by changes at the end of the active period. Fragmented activity later in the active period possibly reflects increasing fatigability and suggests that, for vulnerable PwMS, maintaining long bouts of physically demanding activities gets progressively harder as fatigue sets in. Our findings are in line with prior studies in older adults (without MS) demonstrating positive correlations between ASTP and fatigability (both self-reported fatigability and performance fatigability during a 6-minute walk test), as well as inverse correlations between ASTP and gait speed.^14,18,19^ In a study from the Baltimore Longitudinal Study of Aging combining accelerometry and metrics of walking energy expenditure based on metabolic analysis, older adults with lower energy capacity and higher energy required for walking had higher ASTP.^13^ It is hypothesized that activity fragmentation may be an early compensatory strategy to maintain physical activity in view of lower energy capacity and higher energy needs for mobility. Activity fragmentation may offer insights into processes beyond efficiency of energy utilization for mobility. Increased activity fragmentation has been associated with frailty and with higher risk of all-cause mortality independent of total physical activity volume.^10-12,16,17^ In a longitudinal study of cognitively intact older adults, higher baseline physical activity fragmentation was associated with steeper declines in memory and visuospatial processing.^38^ In another study from the Baltimore Longitudinal Study of Aging, lower activity fragmentation was associated with higher temporal white matter volumes.^39^ Furthermore, higher activity fragmentation has been reported in people with dementia compared to controls, with even higher levels in those with dementia who had lower cognitive scores.^40,41^ Taken together, these findings suggest that activity fragmentation may be closely linked to occult neurodegeneration. In this context, worsening activity fragmentation has been proposed as a potential phenotype of accelerated biological aging. While the exact pathophysiology of this association remains to be elucidated, several plausible mechanisms have been hypothesized including mitochondrial dysfunction with decline in energy reserves and chronic inflammation (“inflammaging”).^42^ Parallels can be drawn between these concepts and MS. In MS, chronic demyelination may predispose to neurodegeneration and, in turn, chronically demyelinated or insufficiently remyelinated axons exhibit conduction delay and failure with prolonged activation, which can manifest clinically as fatigability. It is therefore conceivable that fatigability, as reflected by worsening activity fragmentation throughout an individual’s active period in the current study, is a risk factor for future disability progression and brain atrophy.

In contrast to ASTP, we did not observe any meaningful associations between SATP and disability outcomes or brain atrophy. SATP refers to interruptions of sedentary periods with activity and is postulated to indicate better cardiometabolic health. Prior literature with regards to the clinical relevance of sedentary state fragmentation is inconclusive with conflicting findings regarding to associations with incident frailty in older adults.^15,17^ In the current work, SATP was not associated with risk of disability progression or greater brain volume preservation over time. There are several potential reasons to explain this observation. Participants with significant cardiac comorbidities were not included in this cohort, for which this association may have been discernible. Additionally, sedentary fragmentation, when considered in isolation, is a rather crude measure and does not examine the duration or intensity of activity during the interruptions in sedentary behavior. It is likely that engaging in exercise and minimizing sedentary time is more meaningful than sedentary fragmentation in itself; examining this distinction was beyond the scope of the current work.

Strengths of this study include the novel design and availability of longitudinal accelerometry data. To date, most accelerometry studies have been cross-sectional or have investigated gross activity metrics. In this study, we examined more subtle behavioral patterns and used trends of activity fragmentation throughout the course of one’s active period as a novel framework to gain potential insights into fatigability. We observed stronger associations between activity fragmentation and neurologic outcomes in within-person models rather than mean/between-person models, suggesting that within-person changes in activity are more informative and accelerometry can potentially provide insights at the individual level.

This study has several limitations that warrant discussion. First, the follow-up period, particularly for MRI outcomes, was relatively short. Nonetheless, while a short study timeframe may bias toward null findings, a substantial proportion of participants experienced confirmed disability progression, and meaningful associations of activity fragmentation with both disability and imaging outcomes were observed. Results regarding changes in activity fragmentation appared to be primarily driven by participants with PMS; consequently, the follow-up duration may have been insufficient to capture changes in participants with RRMS. Secondly, our study may have been susceptible to Hawthorne effect, where participants may modify their habitual physical activity behaviors because they are aware they are being monitored with an accelerometer. To minimize this risk, features that display activity levels were disabled; only the time of day was displayed on the screen of the wearable device. Lastly, although metrics of activity fragmentation provide insights into more nuanced activity patterns compared to traditional summary measures, they do not fully capture the granular aspects of mobility. Nonetheless, our findings suggest that increasing activity fragmentation over the course of one’s active period may reflect fatigability and could serve as a risk factor for future disability progression and brain atrophy. Therefore, it may be useful for future studies to explore more precise methods to capture movement patterns and further explore fatigability as a proxy for demyelination-related conduction deficits and reduced physiologic reserve.

In conclusion, we found that PwMS who exhibit increasingly disrupted activity patterns throughout their active period are at a higher risk of neurologic decline, including disability progression during follow-up and accelerated brain atrophy. Monitoring patterns of activity fragmentation could therefore provide valuable insights into subtle physiologic changes and help identify PwMS at risk for accelerated disability accumulation. An essential next step for this line of research is to capture mobility with greater granularity and establish clinically meaningful thresholds for change in PwMS. Regardless, our work underscores that accelerometry-based monitoring is a promising tool for tracking MS progression and may have potential applications both in clinical care and in clinical trials.

## Data Availability

All data produced in the present study are available upon reasonable request to the authors with proper inter-institutional data sharing agreements in place.

## Figure legends

**Supplementary Figure 1.**
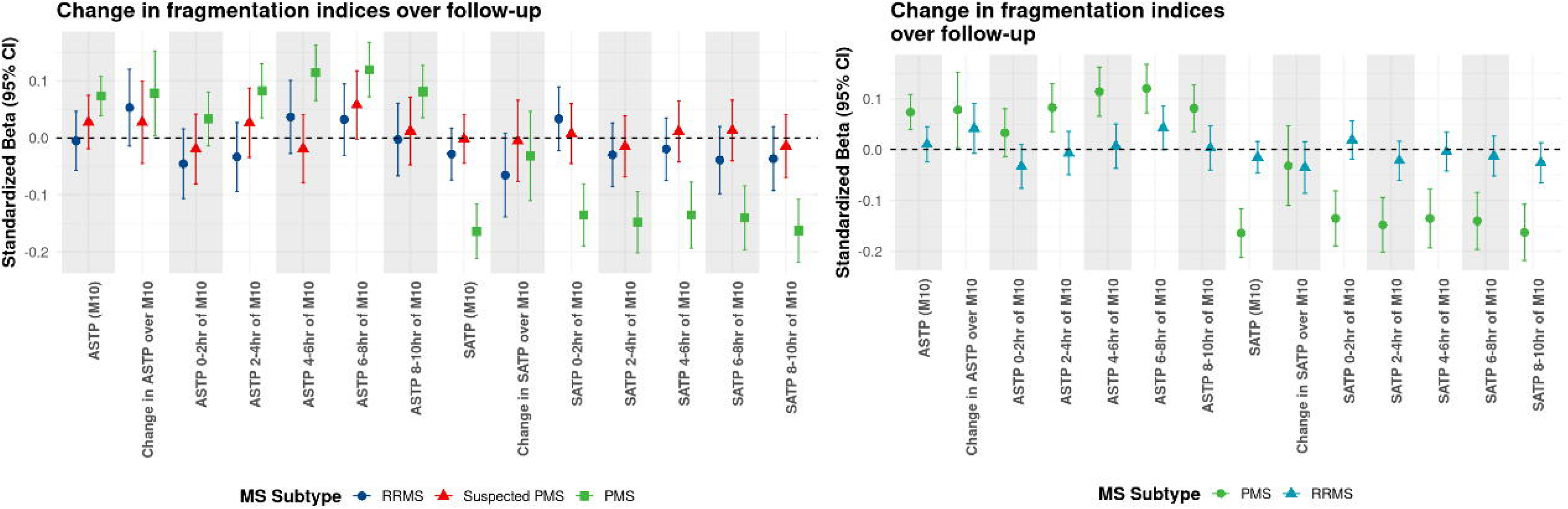
Change in metrics of activity fragmentation during follow-up after adjusting for season. *Estimates are calculated from linear mixed effects models adjusted for age, sex, race and body mass index (BMI). Abbreviations: RRMS: relapsing-remitting MS, PMS: progressive MS, ASTP: active-to-sedentary transition probability, SATP: sedentary-to-active transition probability; M10: most active 10 hours of the day*

**Supplementary Figure 2.**
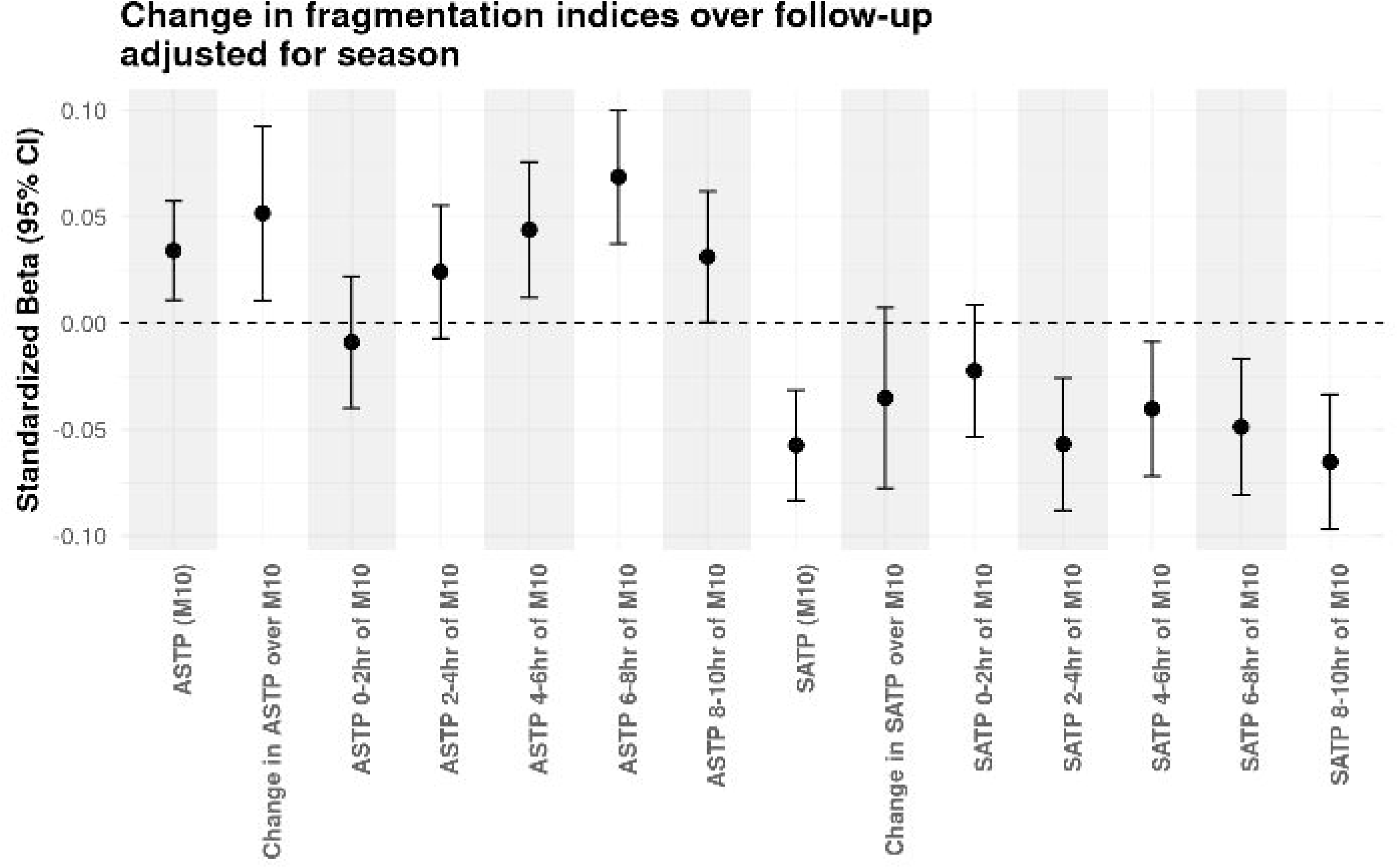
Change in metrics of activity fragmentation by HEAL-MS subgroups (stable RRMS, RRMS with suspected progression/suspected PMS, PMS) and by clinical MS subtype (RRMS, PMS). *Estimates are calculated from linear mixed effects models adjusted for age, sex, race and body mass index (BMI). Abbreviations: RRMS: relapsing-remitting MS, PMS: progressive MS, ASTP: active-to-sedentary transition probability, SATP: sedentary-to-active transition probability; M10: most active 10 hours of the day*

## Tables

**Supplementary Table 1.** Association of metrics of activity fragmentation with risk of EDSS-plus disability worsening during follow-up in sensitivity analyses adjusted for total activity count.

| Variable | Confirmed EDSS-plus progression |  |  |  |
| --- | --- | --- | --- | --- |
|  | Mean/between-person model |  | Within-person model |  |
|  | HR (95 % CI) | P-value | HR (95 % CI) | P-value |
| ASTP (M10) | 0.98 (0.68, 1.40) | 0.9 | 1.12 (0.94, 1.34) | 0.21 |
| Change in ASTP over M10 | 0.95 (0.78, 1.16) | 0.59 | **1.22 (1.01, 1.46)** | **0.03** |
| ASTP 0-2hr of M10 | 0.91 (0.68, 1.21) | 0.5 | 0.92 (0.76, 1.11) | 0.38 |
| ASTP 2-4hr of M10 | 0.91 (0.69, 1.20) | 0.52 | 1.15 (0.95, 1.39) | 0.15 |
| ASTP 4-6hr of M10 | 0.95 (0.71, 1.28) | 0.75 | 1.06 (0.88, 1.28) | 0.52 |
| ASTP 6-8hr of M10 | 0.79 (0.59, 1.07) | 0.13 | 1.08 (0.90, 1.31) | 0.42 |
| ASTP 8-10hr of M10 | 1.00 (0.75, 1.34) | 0.99 | **1.23 (1.04, 1.45)** | **0.02** |
| SATP (M10) | 1.00 (0.75, 1.33) | 0.98 | 0.98 (0.80, 1.20) | 0.84 |
| Change in SATP over M10 | 1.06 (0.88, 1.28) | 0.55 | 0.98 (0.80, 1.19) | 0.81 |
| SATP 0-2hr of M10 | 0.92 (0.71, 1.21) | 0.56 | 0.94 (0.77, 1.14) | 0.51 |
| SATP 2-4hr of M10 | 1.01 (0.77, 1.32) | 0.97 | 1.01 (0.82, 1.24) | 0.93 |
| SATP 4-6hr of M10 | 1.01 (0.78, 1.32) | 0.94 | 1.09 (0.89, 1.33) | 0.42 |
| SATP 6-8hr of M10 | 1.02 (0.78, 1.33) | 0.91 | 1.01 (0.83, 1.23) | 0.9 |
| SATP 8-10hr of M10 | 0.99 (0.75, 1.31) | 0.96 | 0.93 (0.76, 1.13) | 0.46 |

EDSS: expanded disability status scale; HR: hazard ratio; ASTP: active-to-sedentary transition probability, SATP: sedentary-to-active transition probability; M10: most active 10 hours of the day

**Supplementary Table 2.**
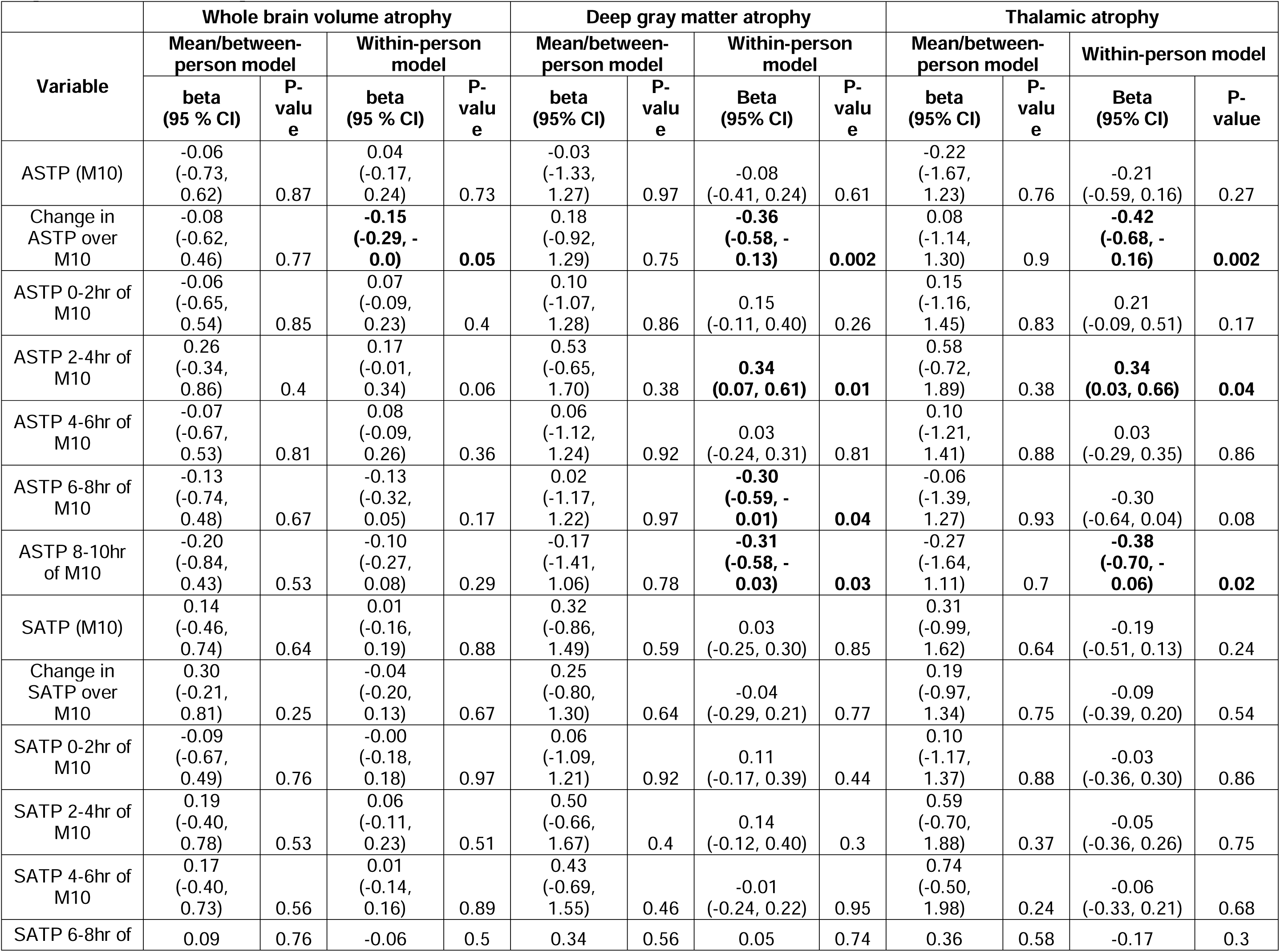
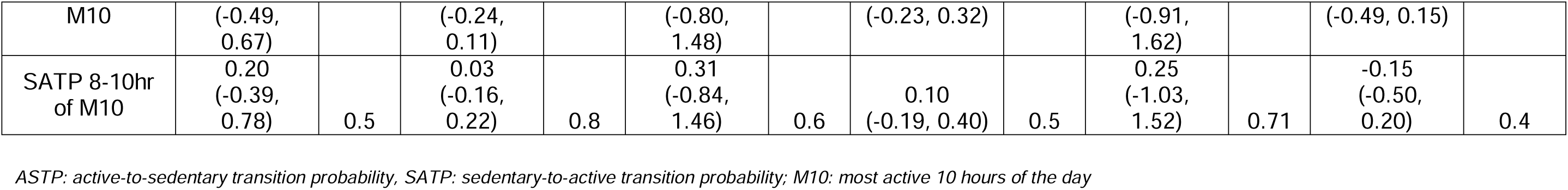
Association of metrics of activity fragmentation with rates of brain compartment atrophy in sensitivity analyses adjusted for total activity count.

